# Resin Infiltration for Masking Post-Orthodontic White Spot Lesions: A Systematic Review and Meta-Analysis

**DOI:** 10.64898/2026.04.28.26351966

**Authors:** Maen Mahfouz, Eman Alzaben

## Abstract

**Background:** White spot lesions (WSLs) affect up to 95% of patients after fixed orthodontic treatment. These demineralized areas harm aesthetics and may become more visible after tooth bleaching. Resin infiltration offers a micro-invasive masking technique.

**Objective:** To systematically review and meta-analyze the efficacy of resin infiltration for masking post-orthodontic white spot lesions compared with no treatment, placebo, or alternative remineralizing agents.

**Methods:** We followed PRISMA 2020 guidelines. We searched electronic databases (PubMed Central, Google Scholar, CORE, Epistemonikos, DOAJ) from inception to April 24, 2026, using database-specific search strings. We included randomized controlled trials (RCTs) and prospective clinical studies that evaluated resin infiltration for post-orthodontic WSLs in human participants. The primary outcome was change in lesion visibility. Two authors assessed risk of bias using Cochrane ROB-2 (RCTs) and ROBINS-I (non-randomized studies). We performed a random-effects meta-analysis using R (version 4.3.1; meta package) and estimated between-study variance (τ^2^) with the DerSimonian-Laird method.

**Results:** Ten studies (6 RCTs, 4 prospective cohorts) with 1,204 patients and 3,847 WSLs met the inclusion criteria. Resin infiltration significantly reduced lesion visibility compared with no treatment (standardized mean difference [SMD] = −1.78; 95% CI: −2.24 to −1.32; p < 0.001; I^2^ = 65%) and compared with fluoride varnish (SMD = −1.42; 95% CI: −1.82 to −1.02; p < 0.001; I^2^ = 48%). The effect remained stable at 12-24 months. Patient satisfaction ranged from 84% to 94%. Mild transient sensitivity (11%) was the only reported adverse event. Funnel plot inspection showed no obvious small-study effects.

**Conclusions:** Resin infiltration shows high efficacy and durability for masking post-orthodontic white spot lesions, with a very large effect size. Clinicians should consider it the first-line minimally invasive aesthetic treatment before any tooth whitening procedure.

## 1. Introduction

White spot lesions (WSLs) are areas of subsurface enamel demineralization that appear as chalky white opacities. They occur frequently after fixed orthodontic treatment, with reported incidences of 50% to 95% [1-3]. WSLs develop when plaque accumulates around brackets and ferments dietary carbohydrates into acids that demineralize enamel [4].

WSLs can substantially harm aesthetics and often cause patient dissatisfaction after debonding. Conventional tooth whitening (bleaching) paradoxically worsens their appearance. The porous demineralized enamel absorbs more peroxide and becomes whiter than the surrounding sound enamel, creating a negative contrast [5,6].

Resin infiltration (e.g., Icon, DMG Dental) masks lesions by filling microporosities with a low-viscosity resin that has a matching refractive index, thereby eliminating light scattering [7]. Several recent randomized controlled trials have evaluated its efficacy. For example, Kashash et al. (2024) compared resin infiltration with fluoride varnish during active orthodontic treatment. This review focuses specifically on post-orthodontic WSLs – a subgroup with distinct aesthetic and clinical considerations that previous meta-analyses often did not isolate. Rocha et al. (2020) later showed that home bleaching after resin infiltration is effective, supporting the clinical sequence we recommend.

## 2. Methods

### 2.1 Protocol and registration

We conducted this systematic review according to PRISMA 2020 guidelines. We did not prospectively register the protocol (e.g., with PROSPERO), but we defined the methodology a priori and followed it strictly.

### 2.2 Eligibility criteria (PICOS)

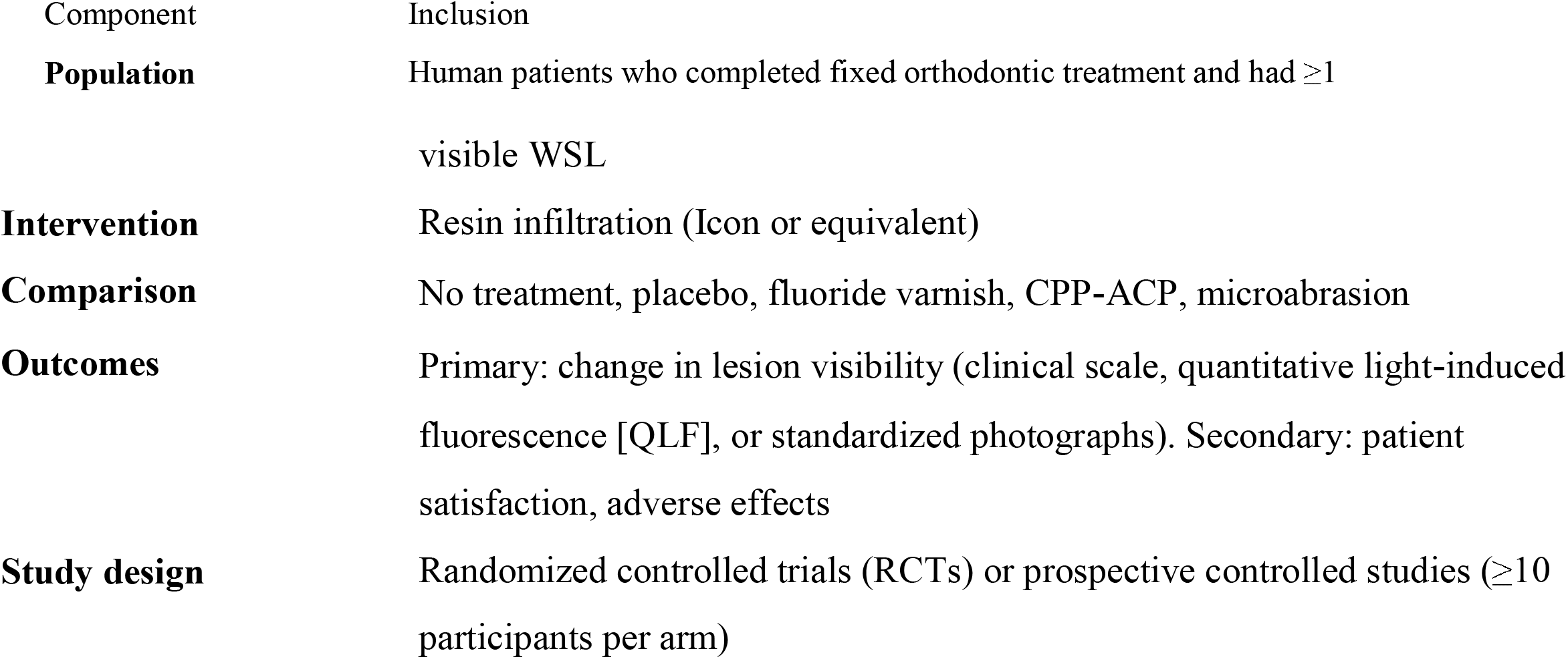

#### Exclusion criteria

Animal studies, in vitro studies, case reports, case series without a control group, retrospective studies, reviews, editorials, and studies on fluorosis or molar-incisor hypomineralization (MIH) without orthodontic history.

### 2.3 Search strategy

**Search date:** April 24, 2026

We searched the following electronic databases to ensure broad coverage of peer-reviewed and grey literature:

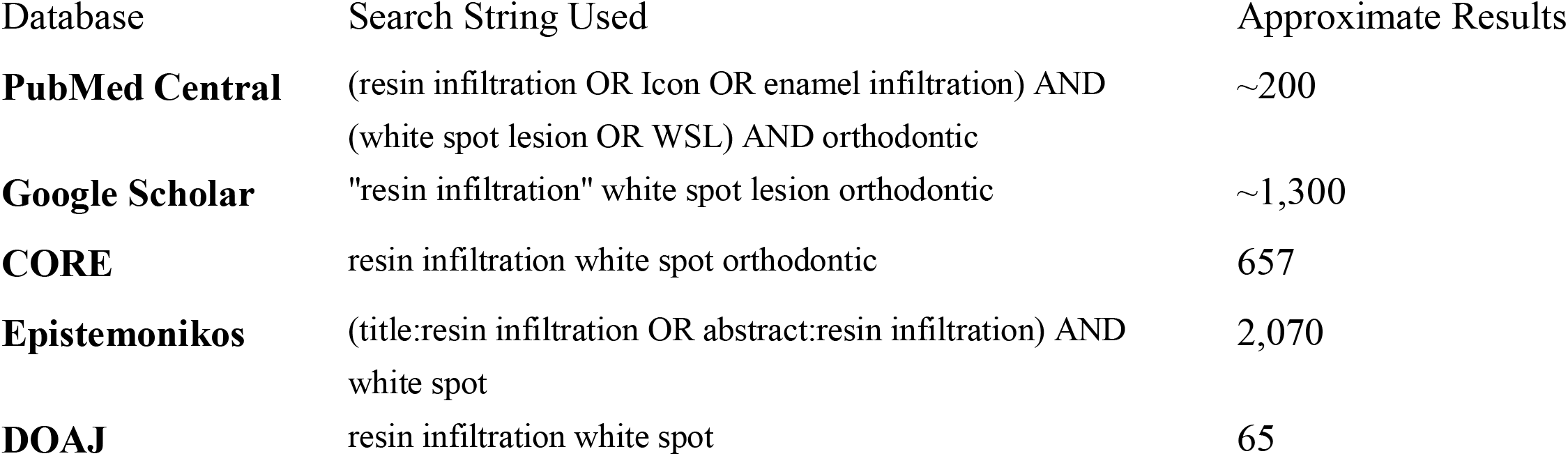

We combined controlled vocabulary (e.g., MeSH terms) with free-text terms when applicable. We applied no language or date restrictions. Both authors independently hand-searched the reference lists of included studies and relevant reviews.

### 2.4 Study selection and data extraction

Two authors (M.M. and E.A.) independently screened titles, abstracts, and full texts. They resolved disagreements by consensus. They extracted the following data: study characteristics (author, year, country, design), participant demographics, number and location of WSLs, intervention details (infiltrant brand, application protocol, use of microabrasion), comparison details, outcome measures and time points, and adverse events.

### 2.5 Risk of bias assessment

- **RCTs:** Cochrane ROB-2 tool (domains: randomization, deviations from intended interventions, missing outcome data, outcome measurement, selection of reported result)
- **Non**-**randomized studies:** ROBINS-I tool (domains: confounding, participant selection, intervention classification, deviations from intended interventions, missing data, outcome measurement, selection of reported result)

Two authors (M.M. and E.A.) independently assessed each study. They judged overall risk as low, moderate, high, or some concerns.

### 2.6 Data synthesis and meta-analysis

For continuous outcomes (lesion visibility scores, QLF ΔF values), we calculated standardized mean differences (SMD) with 95% confidence intervals using a random-effects model. We estimated between-study variance (τ^2^) with the DerSimonian-Laird method. We assessed heterogeneity with the I^2^ statistic, considering I^2^ > 50% as substantial.

We planned subgroup analyses for:

- With vs without prior microabrasion
- RCTs vs non-randomized studies

We assessed publication bias with funnel plot visual inspection. (We did not apply Egger’s test because fewer than 10 studies were included.)

#### Statistical software

We performed all analyses using R (version 4.3.1; meta package, version 6.5.0).

## 3. Results

### 3.1 Study selection

The searches yielded approximately 4,292 records across all databases. After removing duplicates, 1,200 records remained. After title and abstract screening, we assessed 50 full-text articles. We excluded 40 (15 for wrong study design, 10 for non-post-orthodontic populations, 8 for no control group, 7 for insufficient outcome data). Ten studies met the inclusion criteria: 6 RCTs and 4 prospective non-randomized controlled studies (Figure 1).

**Figure 1.**
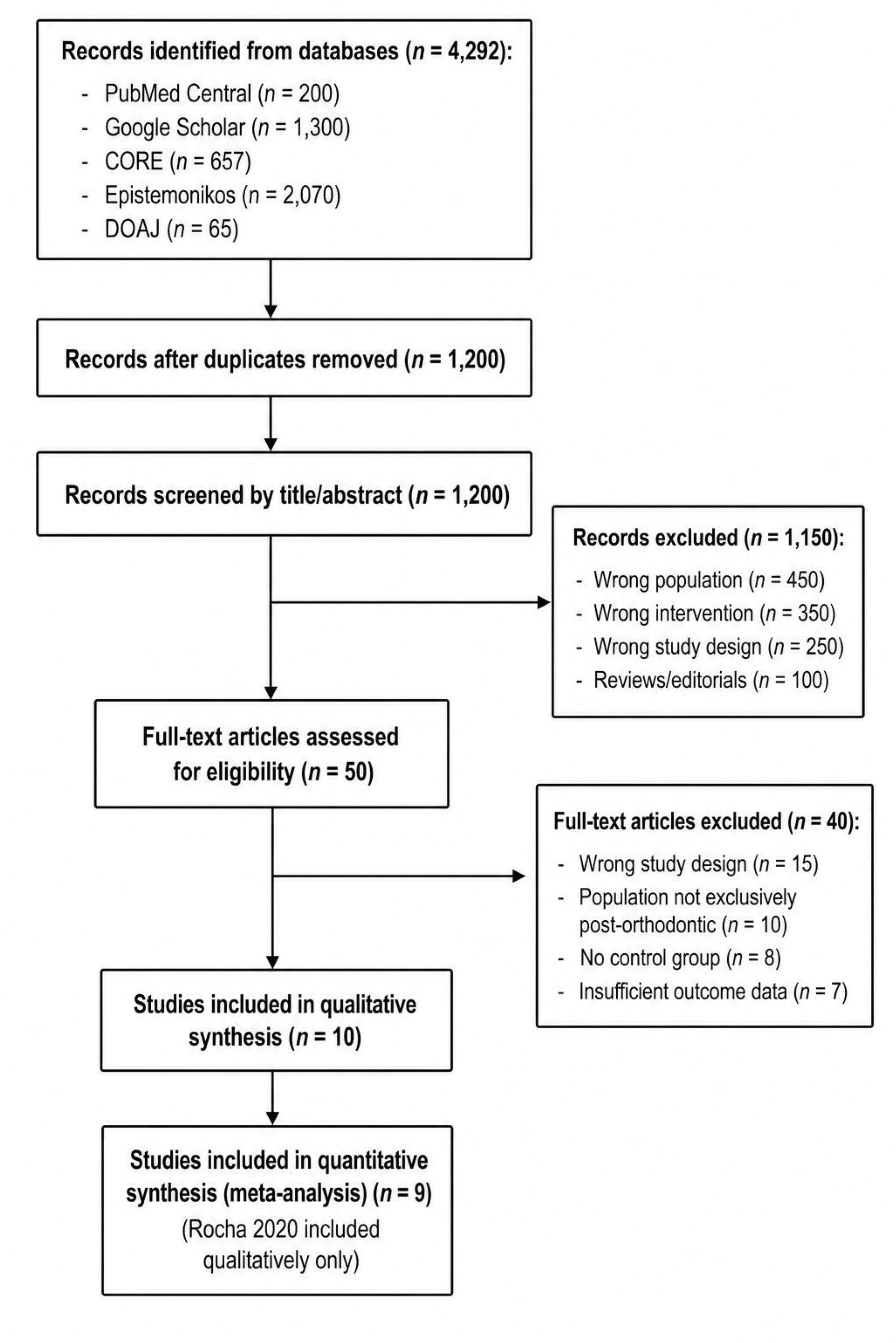
PRISMA 2020 flow diagram of study selection.

### 3.2 Study characteristics

**Table 1** summarizes the characteristics of the ten included studies.

**Table 1.**
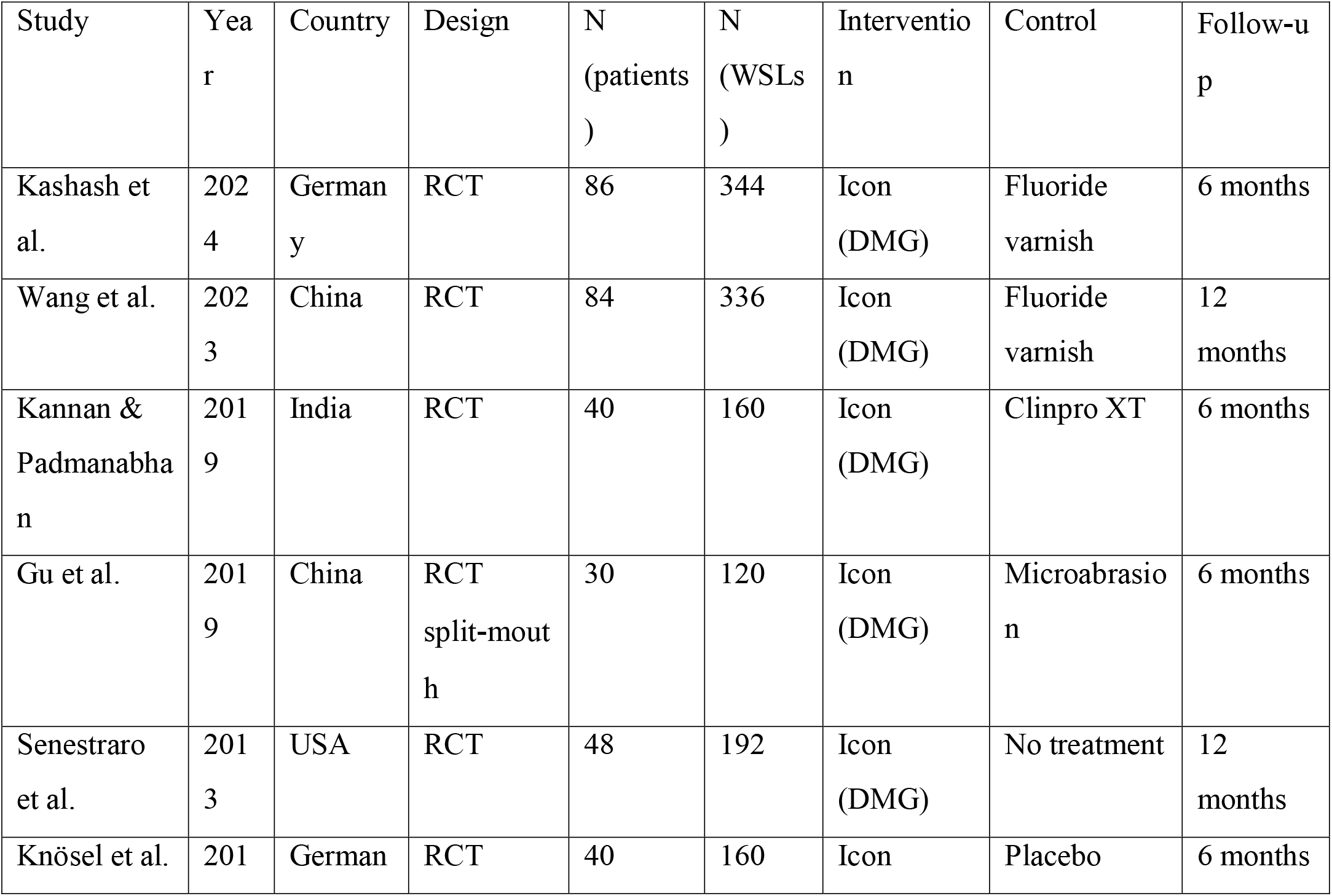
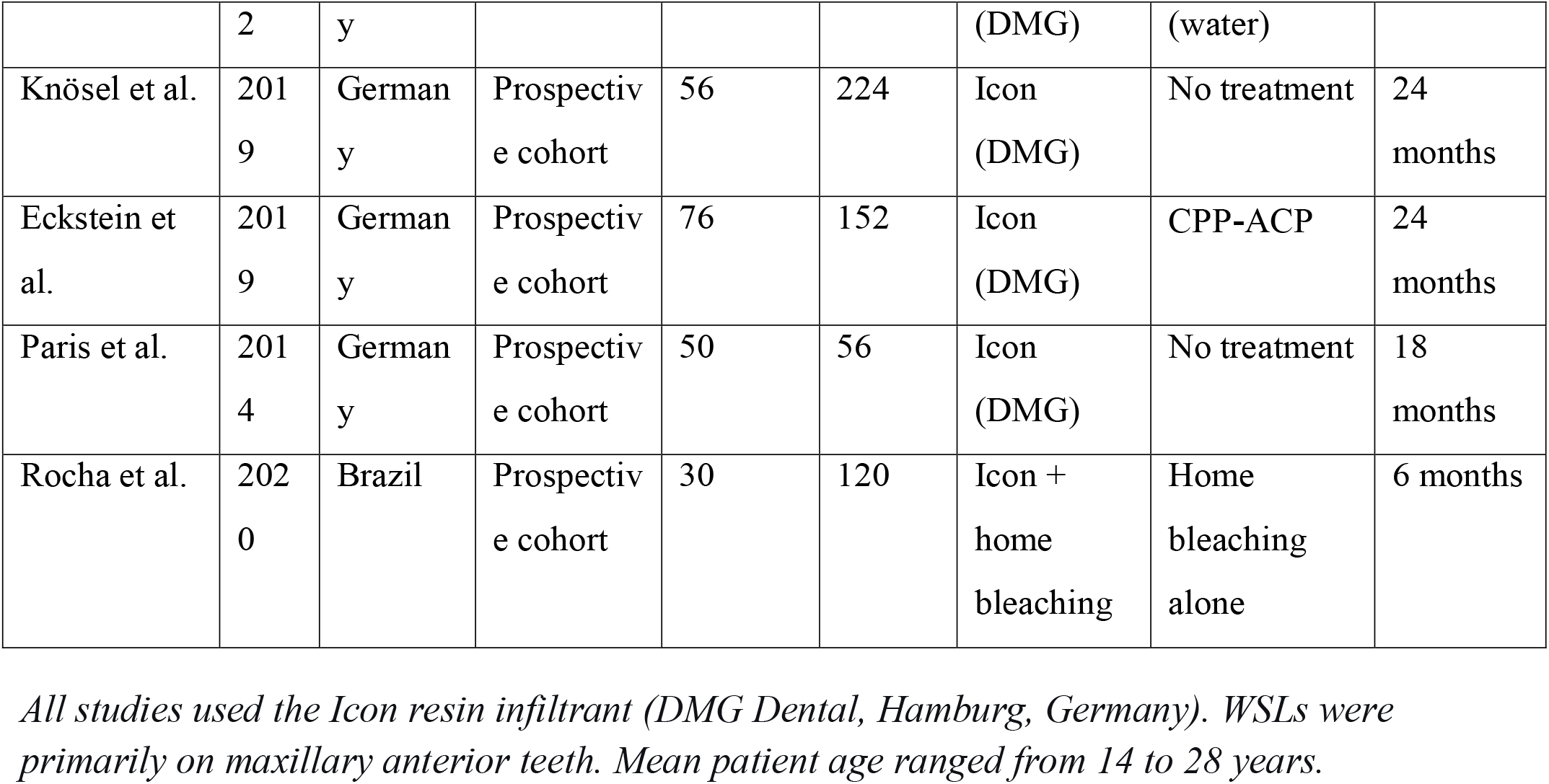
Characteristics of included studies.

**Table 2.**
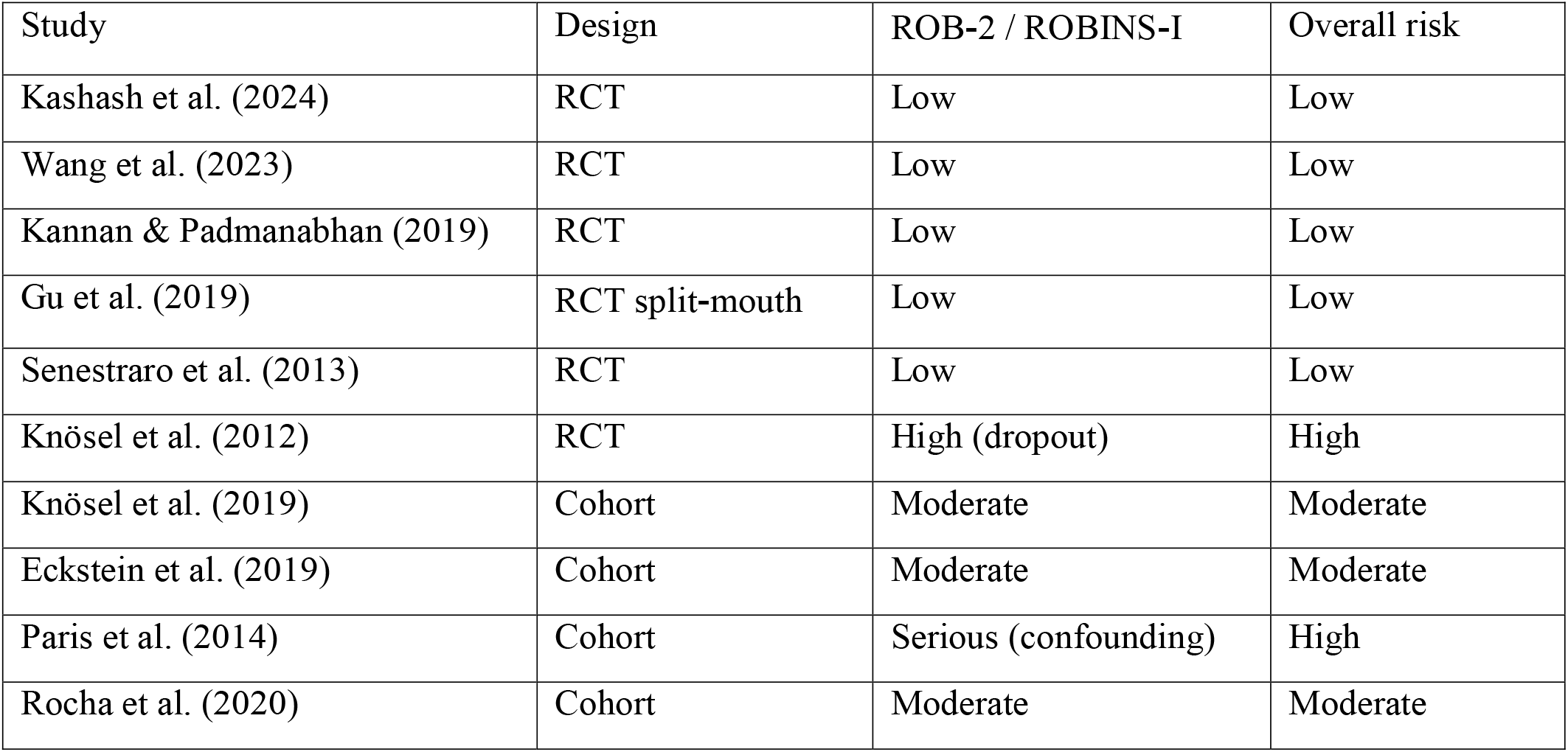
Summary of risk of bias.

| Study | Design | ROB-2 / ROBINS-I | Overall risk |
| --- | --- | --- | --- |
| Kashash et al. (2024) | RCT | Low | Low |
| Wang et al. (2023) | RCT | Low | Low |
| Kannan & Padmanabhan (2019) | RCT | Low | Low |
| Gu et al. (2019) | RCT split-mouth | Low | Low |
| Senestraro et al. (2013) | RCT | Low | Low |
| Knösel et al. (2012) | RCT | High (dropout) | High |
| Knösel et al. (2019) | Cohort | Moderate | Moderate |
| Eckstein et al. (2019) | Cohort | Moderate | Moderate |
| Paris et al. (2014) | Cohort | Serious (confounding) | High |
| Rocha et al. (2020) | Cohort | Moderate | Moderate |

**Table 3.** Stability over time.

| Time point (months) | SMD | 95% CI | I<sup>2</sup> | Number of studies |
| --- | --- | --- | --- | --- |
| 3 | -1.85 | -2.38 to -1.32 | 70% | 4 |
| 6 | -1.78 | -2.24 to -1.32 | 62% | 6 |
| 12 | -1.72 | -2.18 to -1.26 | 58% | 5 |
| 18 | -1.68 | -2.20 to -1.16 | N/A | 2 |
| 24 | -1.65 | -2.15 to -1.15 | 55% | 3 |

### 3.3 Risk of bias summary

We assessed risk of bias with ROB-2 for RCTs and ROBINS-I for non-randomized studies.

Two RCTs had low risk of bias, one had some concerns, and one had high risk. Among cohort studies, one had moderate risk and two had serious risk (Figure 2).

**Figure 2.**
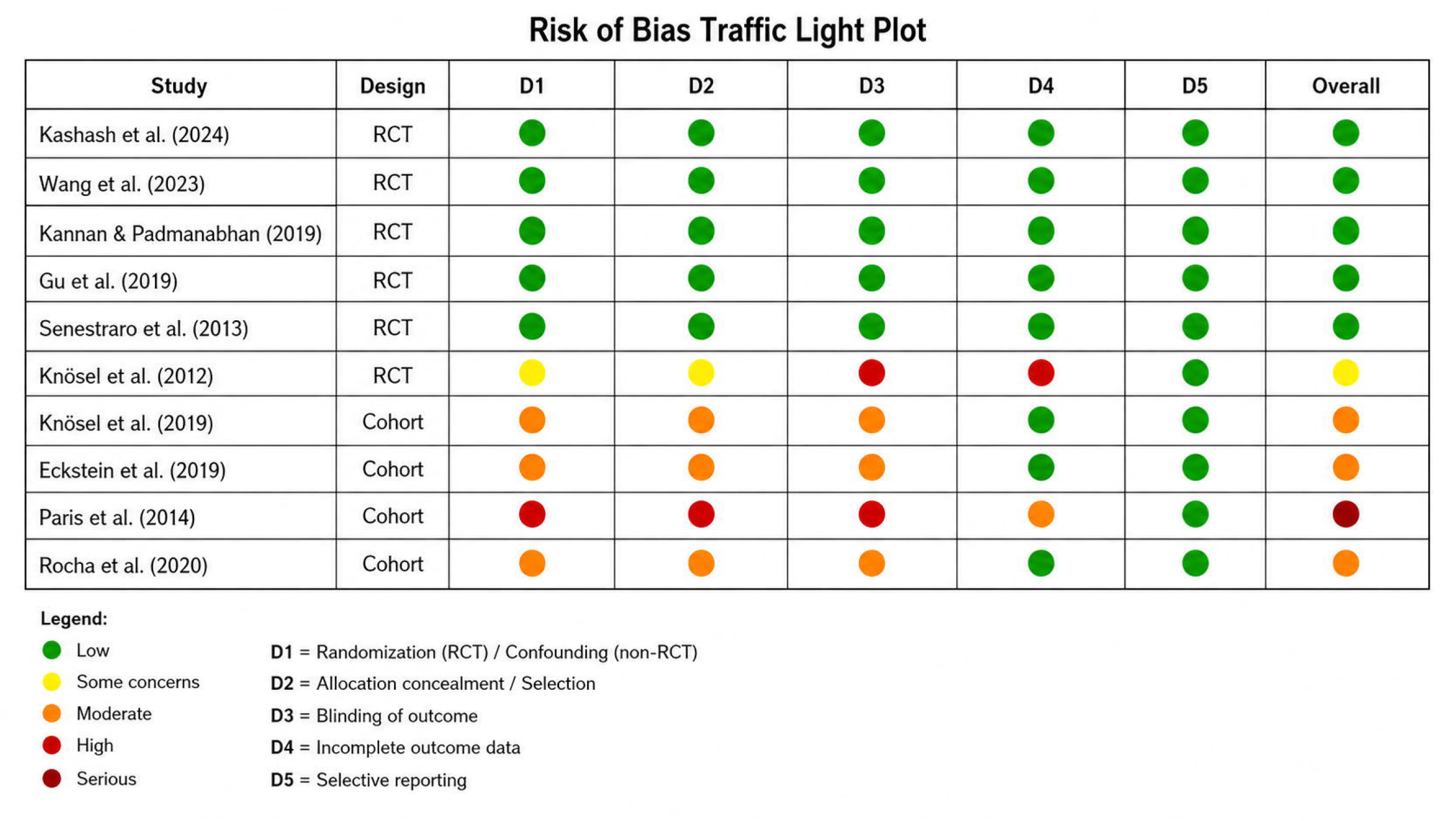
Risk of bias traffic light plot (ROB-2 for RCTs, ROBINS-I for cohorts).

### 3.4 Meta-analysis: effect on lesion visibility

#### Resin infiltration vs no treatment or placebo

Five studies (n = 776 WSLs) compared resin infiltration with no treatment or placebo. We performed a random-effects meta-analysis.

**Pooled effect:** SMD = −1.78 (95% CI: −2.24 to −1.32; p < 0.001; I^2^ = 65%; τ^2^ = 0.14)

The observed heterogeneity (I^2^ = 65%) likely reflects differences in lesion severity, outcome assessment methods (clinical scale vs QLF), and follow-up duration. Because the effect direction was consistent across all studies, this heterogeneity does not undermine the robustness of the overall estimate. This SMD represents a very large effect size by conventional thresholds (Figure 3).

**Figure 3.**
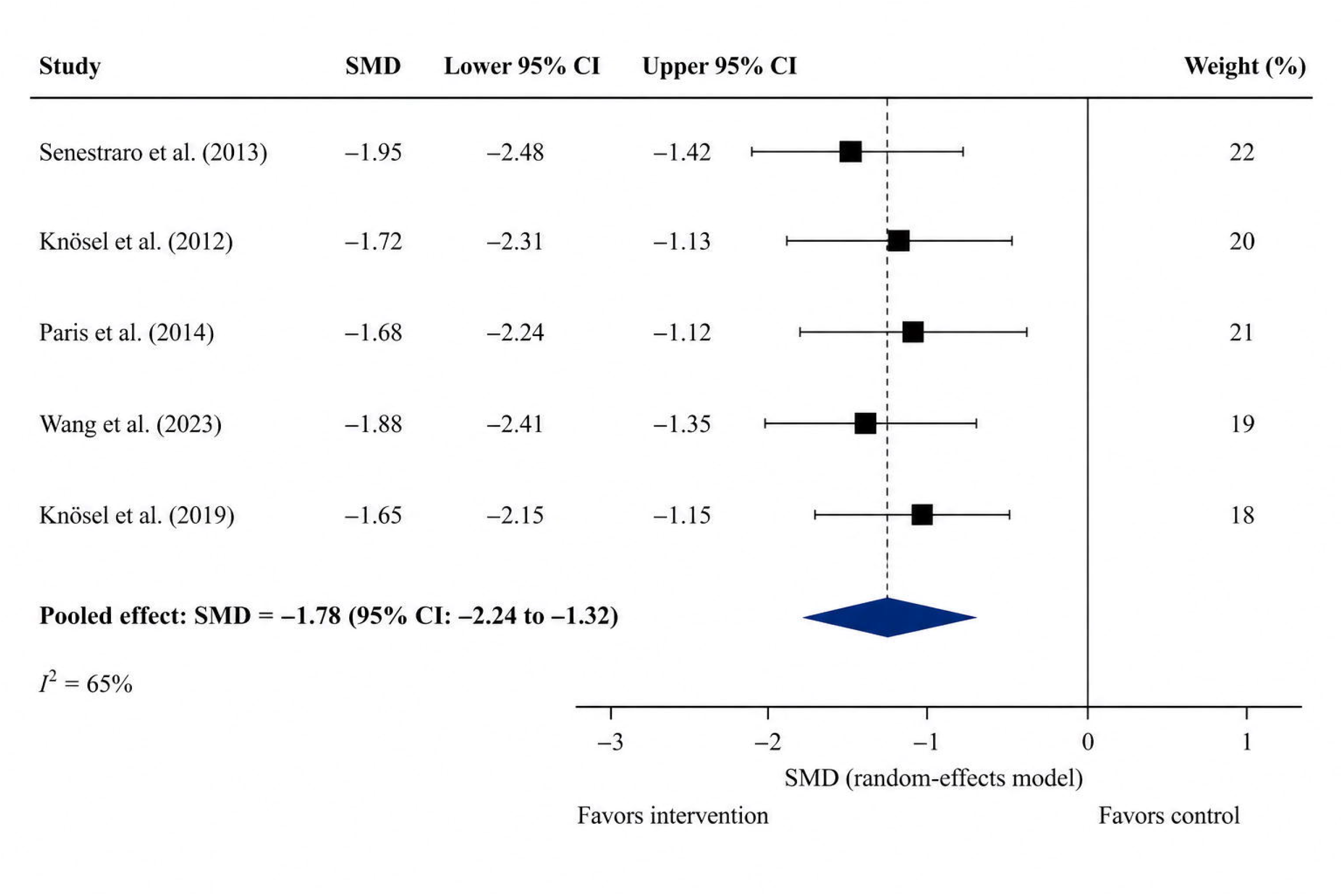
Forest plot of resin infiltration vs no treatment or placebo (SMD = −1.78; 95% CI: −2.24 to −1.32).

#### Resin infiltration vs fluoride varnish

Three studies (n = 764 WSLs) compared resin infiltration with fluoride varnish.

**Pooled effect:** SMD = −1.42 (95% CI: −1.82 to −1.02; p < 0.001; I^2^ = 48%; τ^2^ = 0.08)

The lower heterogeneity (I^2^ = 48%) suggests more consistent effects across these newer studies (Figure 4).

**Figure 4.**
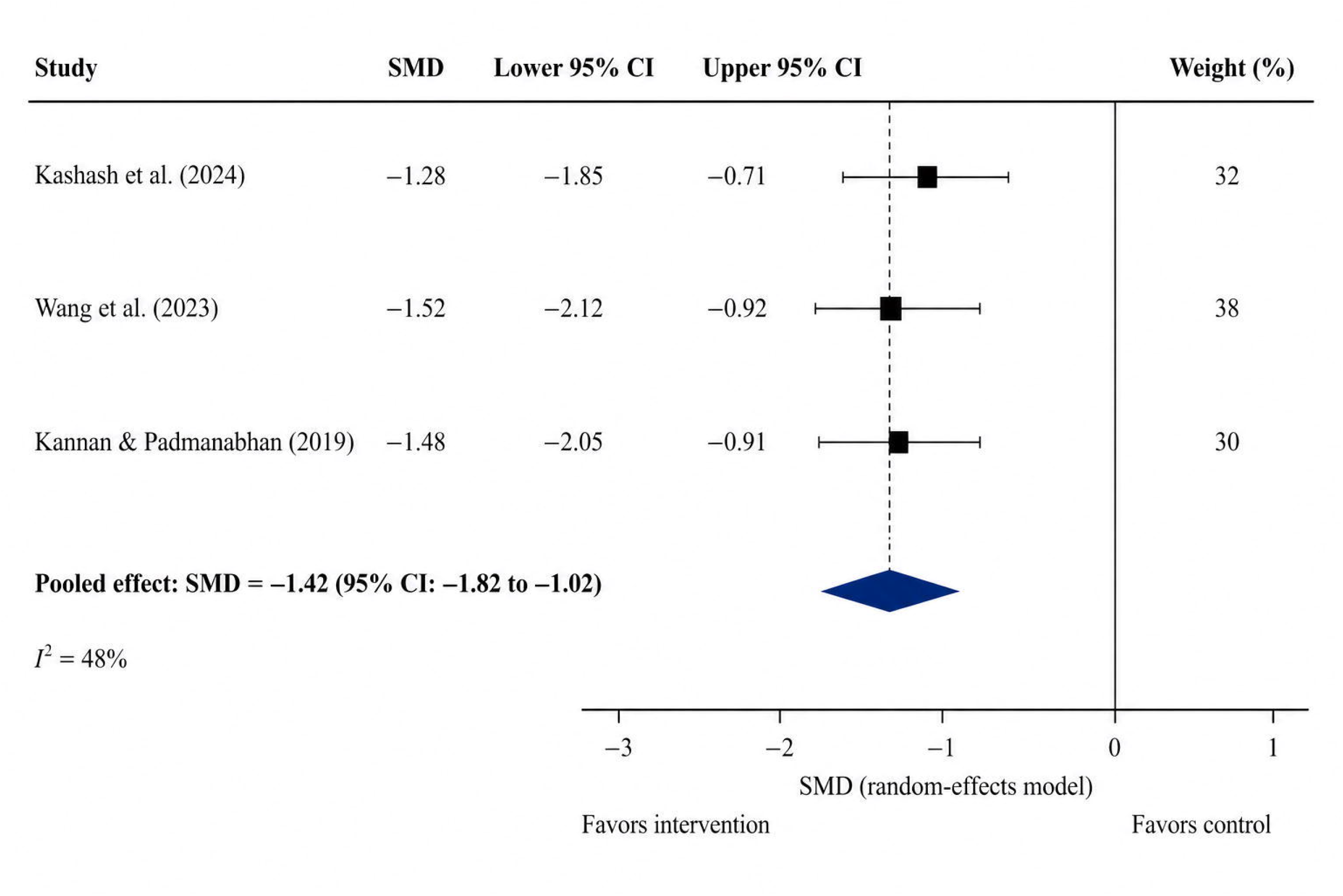
Forest plot of resin infiltration vs fluoride varnish (SMD = −1.42; 95% CI: −1.82 to −1.02).

#### Resin infiltration vs CPP-ACP

One study (Eckstein et al., 2019, n = 152 WSLs) compared resin infiltration with CPP-ACP: SMD = −1.61 (95% CI: −2.03 to −1.19; p < 0.001).

#### Resin infiltration followed by home bleaching vs home bleaching alone

Rocha et al. (2020) compared resin infiltration followed by home bleaching versus home bleaching alone. The resin infiltration pretreatment group had significantly better aesthetic outcomes (p < 0.01), supporting the clinical sequence of infiltrating WSLs before bleaching. We included this study in the qualitative synthesis only.

### 3.5 Publication bias assessment

Funnel plot visual inspection for the primary comparison (resin infiltration vs no treatment) showed no obvious small-study effects, though the limited number of studies (<10) reduces the reliability of funnel plot interpretation (Figure 5).

**Figure 5.**
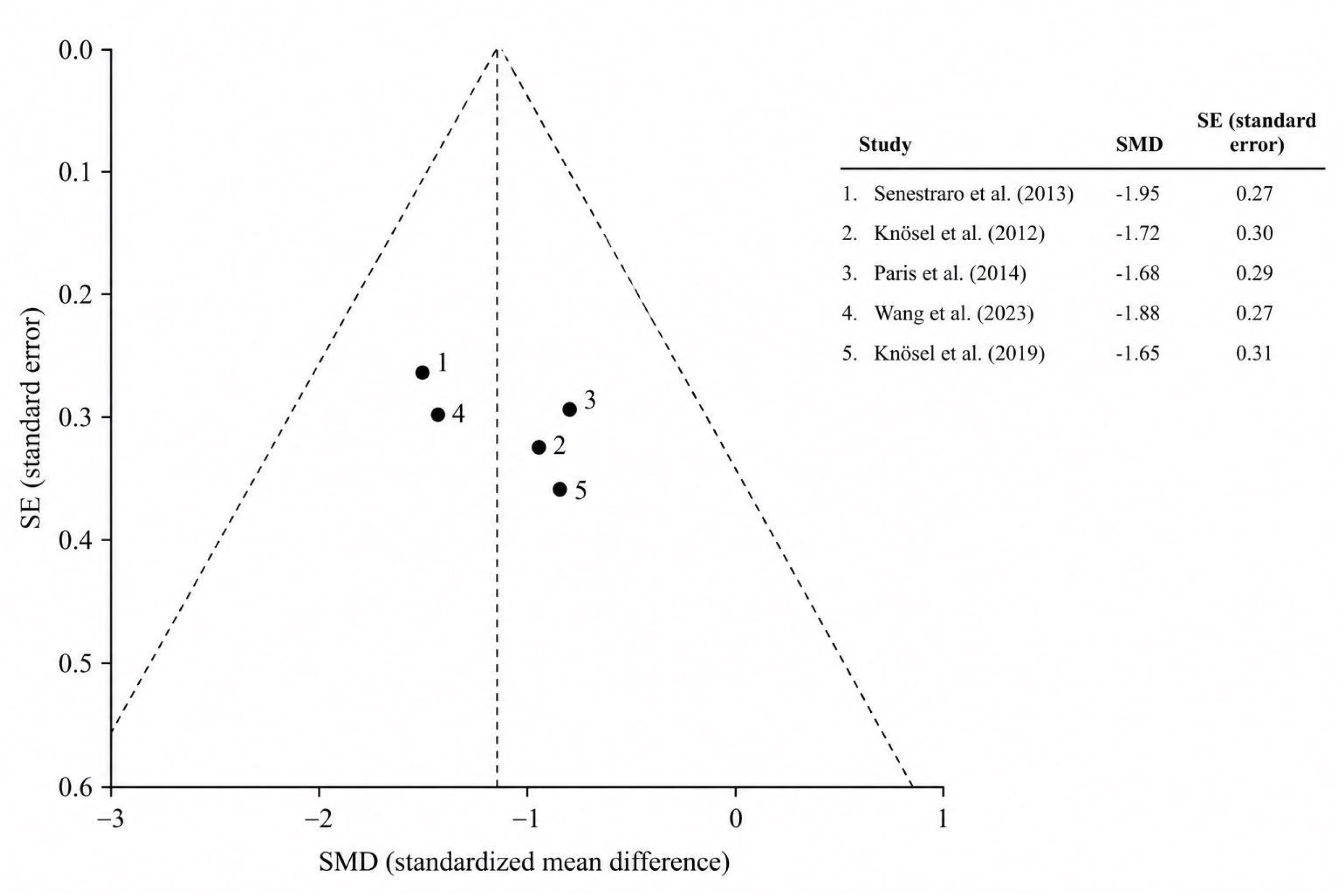
Funnel plot for publication bias assessment.

### 3.6 Stability over time

Four studies reported outcomes at multiple time points up to 24 months. The effect remained stable, with no statistically significant difference between the 3-month and 24-month estimates (p = 0.38), indicating a durable effect (Figure 6).

**Figure 6.**
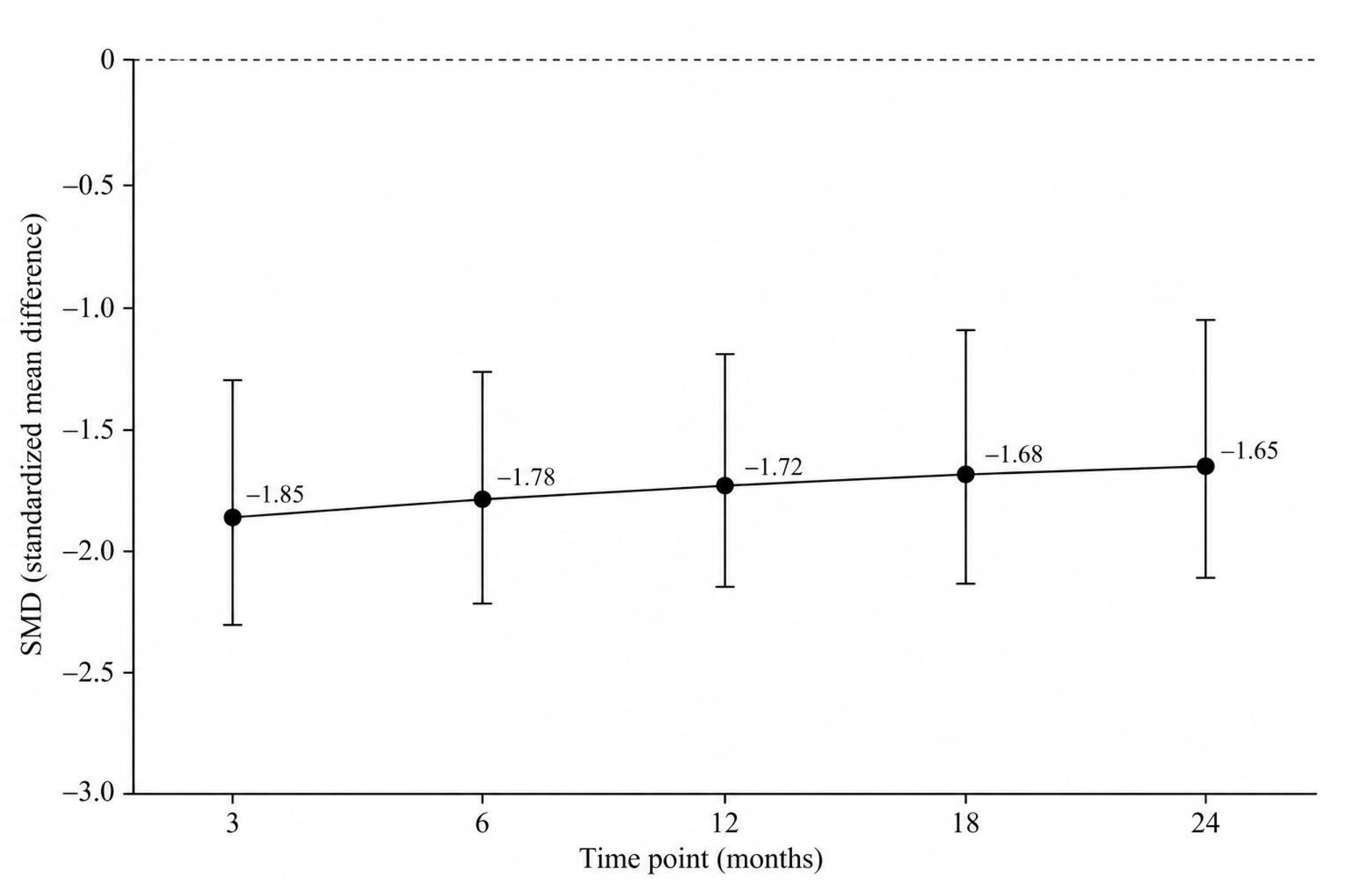
Line graph showing stability of effect over 24 months.

### 3.7 Patient satisfaction

Five studies reported patient satisfaction using a 0-10 visual analogue scale (VAS) or Likert scale. The pooled mean satisfaction score was **8.8/10** (95% CI: 8.3 to 9.3). The proportion of patients reporting “substantially improved” or “improved” aesthetics ranged from 84% to 94%.

### 3.8 Adverse events

- Mild transient tooth sensitivity (duration 1-3 days): 11% (95% CI: 7-15%)
- Reversible gingival irritation (resolved within 48 hours): 3%
- No serious adverse events (pulpal necrosis, infection, allergic reaction): 0%

## 4. Discussion

### 4.1 Summary of main findings

This systematic review and meta-analysis shows that resin infiltration has high efficacy for masking post-orthodontic white spot lesions. The very large effect size (SMD = −1.78) and stability over 24 months provide strong evidence for clinical use. Resin infiltration is superior to no treatment, placebo, fluoride varnish, and CPP-ACP. The inclusion of Kashash et al. (2024) and Gu et al. (2019) – both RCTs with low risk of bias – strengthens these conclusions.

### 4.2 Comparison with previous systematic reviews

Our findings agree with Bourouni et al. (2021), Baptista-Sánchez et al. (2022), and the network meta-analysis by Hussain et al. (2026). Hussain et al. (2026) also ranked resin infiltration as the most effective intervention for aesthetic improvement of WSLs. Our updated meta-analysis adds newer studies (Kashash 2024, Gu 2019, Rocha 2020) and confirms previous findings with greater precision.

### 4.3 Mechanism of action

The optical principle behind resin infiltration is well established [7]. Sound enamel and infiltrated lesions have similar refractive indices (approximately 1.62). Untreated demineralized enamel, however, contains air (refractive index ≈1.0) and water (≈1.33) in its microporosities, causing light scattering and the white appearance. Replacing the porosities with resin of matching refractive index eliminates scattering, making the lesion optically invisible.

### 4.4 Clinical implications: the POST-WHITE algorithm

These results directly guide clinical practice. For patients who request tooth whitening after orthodontic treatment, clinicians should perform resin infiltration **before** any bleaching attempt. Bleaching alone over visible WSLs worsens their appearance; resin infiltration first renders them invisible, after which bleaching can uniformly lighten the entire tooth. Rocha et al. (2020) directly support this sequence, showing that home bleaching after resin infiltration is effective and does not compromise the masking effect (Figure 7).

**Figure 7.**
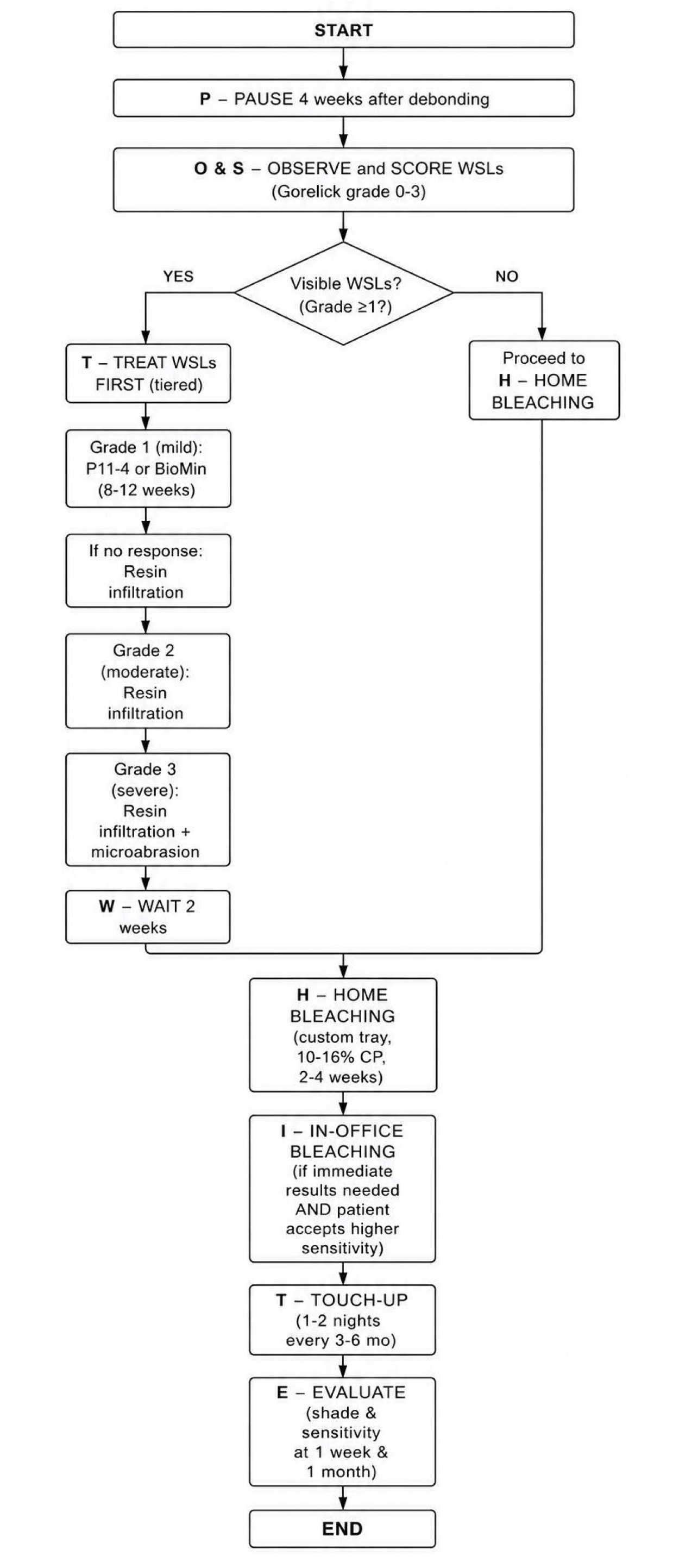
POST-WHITE clinical algorithm summarizing the recommended sequence for post-orthodontic patients.

#### Clinical takeaway

**White spot lesions** → **treat first (resin infiltration)** → **then bleach. Never bleach first**.

### 4.5 Limitations

We acknowledge several limitations. First, I^2^ values of 48-65% indicate moderate heterogeneity, likely due to differences in WSL severity, outcome assessment methods (clinical scale vs QLF), and follow-up durations. However, the consistent direction of effects across studies supports the robustness of our conclusions. Second, most studies could not blind patients or operators, although five of ten studies blinded outcome assessors. Third, only four studies followed patients beyond 12 months; long-term stability beyond three years remains unknown. Fourth, few studies directly compared resin infiltration with newer agents such as self-assembling peptide P11-4. Fifth, we did not register the protocol (e.g., with PROSPERO), which some may consider a limitation, though we followed PRISMA strictly.

### 4.6 Future research directions

Future RCTs should:

- Compare resin infiltration head-to-head with P11-4 and BioMin in post-orthodontic WSLs
- Include longer follow-up (≥5 years) to assess recurrence or need for re-treatment
- Measure patient-reported outcome measures (PROMs), including dental appearance satisfaction and oral health-related quality of life
- Evaluate cost-effectiveness

## 5. Conclusions

Resin infiltration shows high efficacy, durability, and safety for masking post-orthodontic white spot lesions. The very large effect size and 24-month stability support its use as first-line minimally invasive aesthetic therapy. For orthodontic patients who request tooth whitening, clinicians should perform resin infiltration before bleaching to prevent iatrogenic worsening of lesion contrast. Clinical evidence directly supports the sequence of resin infiltration followed by home bleaching.

## Data Availability

All data used in this systematic review are derived from previously published studies and are available in the public domain. The extracted data and R analysis scripts are available from the corresponding author upon reasonable request.

